# VOGeo-Gaze: Calibration-Free, Geometry-Aware Deep Learning for Real-Time Gaze Tracking in Clinical Video-Oculography

**DOI:** 10.64898/2026.05.27.26354254

**Authors:** Jingkang Zhao, Seyed-Ahmad Ahmadi, Julian Decker, Peter zu Eulenburg, Andreas Zwergal, Virginia L. Flanagin, Max Wuehr

**Affiliations:** German Center for Vertigo and Balance Disorders (DSGZ), LMU University Hospital, Munich, Germany; Graduate School of Systemic Neurosciences, Ludwig Maximilian University Munich, Munich, Germany; NVIDIA, Munich, Germany; Schön Clinic Bad Aibling, Bad Aibling, Germany; Department of Neurology, LMU University Hospital, Munich, Germany; Department of Neuroradiology, LMU University Hospital, Munich, Germany

**Keywords:** Clinical gaze tracking, Video-oculography, Calibration-free, Device-independent, Geometry-aware

## Abstract

Quantitative eye movement analysis is important for neurological diagnostics, yet existing video-oculography (VOG) systems typically require calibration, device-specific settings, or accurate gaze labels. We present VOGeo-Gaze, a real-time, calibration-free, geometry-aware neural network that estimates gaze by reconstructing anatomically meaningful eyeball parameters from image features. The method combines segmentation-driven projection geometry, a refraction-aware pupil correction module, and temporal anatomical stabilization, so gaze is derived from interpretable eye geometry rather than direct angular regression. Trained only on the public TEyeD dataset with weak gaze supervision, VOGeo-Gaze was evaluated on 116 clinical recordings from 17 patients and 19 healthy subjects using EyeSeeCam, a clinical gold-standard VOG system. It achieved median absolute angular errors of 0.33° horizontally and 0.35° vertically, with nearly 92% of recordings below 1° error while operating at >300 FPS. These results demonstrate sub-degree clinical gaze estimation without subject-specific calibration, camera intrinsics, or accurate gaze labels, providing a scalable and interpretable alternative to conventional VOG pipelines. Code is available at https://github.com/DSGZ-MotionLab/VOGeo-Gaze.

## 1 Introduction

Head-mounted video-oculography (VOG) systems provide eye-in-head kinematics and are routinely used in clinical neuro-otology. Infrared-based systems such as EyeSeeCam integrate high-speed pupil imaging with inertial sensing and a manufacturer-calibrated geometric pipeline, and are validated for vestibular testing [16,2].

Fig. 1 illustrates three representative paradigms for VOG gaze estimation. Traditional analytical geometric methods (Fig. 1a) estimate horizontal and vertical gaze angles (*θ*_*H*_, *θ*_*V*_) through calibration-bound geometric reconstruction, typically based on pupil–corneal reflection (PCCR) geometry, as implemented in systems such as EyeSeeCam, which achieve high angular precision but rely on fixed hardware configurations and subject-specific calibration [9,16]. Glint-free model-based approaches, initiated by Swirski et al. and extended by Dierkes et al. with refraction-aware eyeball modeling, as well as following learning-assisted methods such as 3DeepVOG, relax explicit intrinsic constraints [17,6,20]. However, their accuracy depends on calibration to stabilize anatomical parameters (e.g., eyeball center and optic axis), limiting robustness under headset misalignment, device slippage, or low-amplitude gaze motion.

**Fig. 1.**
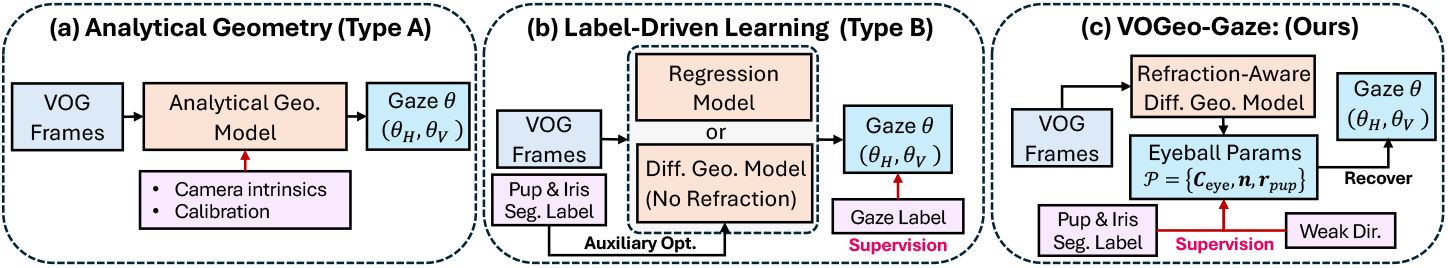
Three paradigms for VOG gaze estimation. (a) Analytical geometry: calibration-dependent closed-form estimation of (*θ*_*H*_, *θ*_*V*_) with known camera intrinsics. (b) Label-driven learning: direct regression or simplified (non-refractive) differentiable models trained with explicit gaze supervision, optimizing in angular space. (c) VOGeo-Gaze (ours): refraction-aware differentiable reconstruction that optimizes anatomical parameters (**c**_eye_, **n**, *r*_pup_) in parameter space via segmentation and weak directional constraints. Gaze is derived without camera intrinsics, subject-specific calibration, or accurate angular labels.

As an alternative paradigm, label-driven learning approaches (Fig. 1b) directly regress gaze angles, such as NVGaze, or incorporate simplified differentiable eye models trained with explicit gaze supervision, including Popovic et al. and De^2^Gaze [11,14,19]. These methods operate primarily in angular space and rely on large-scale, accurate gaze annotations. However, public datasets such as TEyeD provide gaze annotations that are derived from image-based segmentation and landmark geometry rather than independent measurement devices, potentially limiting the achievable clinical accuracy [7].

To address these limitations, we propose VOGeo-Gaze, a calibration-free geometry-constrained framework that shifts optimization from angular space to anatomical parameter space (Fig. 1c). Instead of directly regressing gaze angles (*θ*_*H*_, *θ*_*V*_), the model reconstructs latent eyeball parameters (**c**_eye_, **n**, *r*_pup_) using a differentiable reformulation of the anatomically accurate, refraction-aware two-sphere eye model of Dierkes et al. [5]. Supervision is segmentation-driven and complemented by weak projection-based directional constraints, removing reliance on camera intrinsics, subject-specific calibration, and accurate gaze labels. The gaze vector is derived from the reconstructed optic axis of the eyeball.

VOGeo-Gaze makes three primary contributions: 1) It introduces a fully differentiable, refraction-aware anatomical eye model that enables physically interpretable parameter reconstruction; differentiability allows segmentation-level supervision to propagate directly into geometric parameter estimates without heuristic post-fitting. 2) It achieves sub-degree clinical gaze accuracy without the need for accurate gaze supervision, by enforcing consistency between projected image features and reconstructed 3D eyeball geometry. 3) It requires no subject-specific calibration or camera intrinsics: given only raw VOG frames, the framework directly recovers anatomical parameters and gaze direction, making the design applicable across devices by construction.

## 2 Methods

### 2.1 VOGeo-Gaze Architecture

VOGeo-Gaze estimates gaze without subject-specific calibration or camera intrinsics by reconstructing eye parameters *𝒫* = {**c**_eye_, **n**, *r*_pup_} (eyeball center, optic axis, pupil radius) from projection geometry 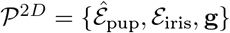 (entrance pupil ellipse, iris ellipse, prior gaze direction) extracted from the image. Throughout,^marks entrance-pupil quantities; ^2*D*^ marks projection; ^*R*^ refraction-derived; ^*^ synthesised ground-truth; subscripts denote component or sample index.

#### Prior Projection Geometry Estimation

Each frame is processed by a lightweight 4-stage ResUNet (channels 16 →128) [13,20] to predict pupil, iris, and eyelid masks. The largest connected components are retained and fitted with ellipses *ℰ* = {*θ*, **c**^2*D*^, *a, b*}, yielding the entrance pupil 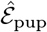 and iris ellipse *ℰ*_iris_ (Fig. 2I(a)).

**Fig. 2.**
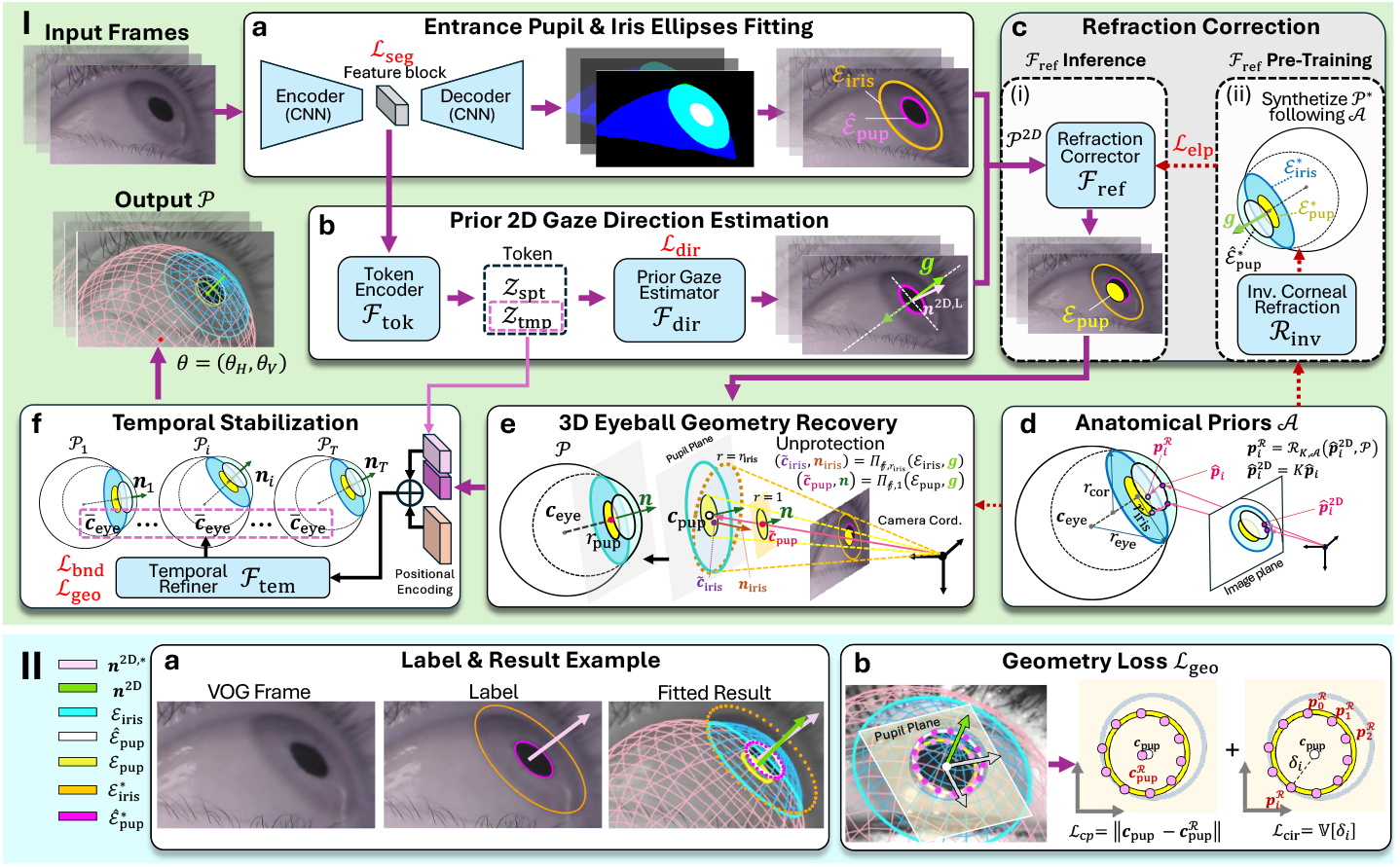
Overview of the VOGeo-Gaze architecture. Panel I: (a) Semantic segmentation and ellipse fitting producing entrance pupil and iris ellipses. (b) Token-based prior gaze estimation resolving minor-axis ambiguity. (c,d) Refraction-aware pupil correction (inference and training) via prior projection geometry and anatomical priors. (d–e) 3D eyeball parameter recovery from prior projection geometry and anatomical priors. (e– f) Temporal stabilization across frames. Panel II: Visualization of the effect from the Geometry Loss.

Encoder bottleneck features are summarized into geometric tokens **Z** ∈ ℝ^256^ by concatenating a 128-D spatial descriptor and a 128-D temporal descriptor (Fig. 2I(b)). We use **Z**_spt_ and **Z**_tmp_ for direction estimation and temporal stabilization, respectively.

A projected gaze prior **g** is approximated by the unit minor axis of the entrance-pupil ellipse 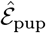, which has a two-fold sign ambiguity. Let 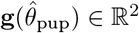 denote the (unsigned) unit minor-axis direction given the fitted ellipse orientation 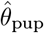. A prior gaze estimator network ℱ_dir_ (hidden dim. 128) predicts a 2D direction from tokens **Z** under weak supervision by projected gaze labels **n**^2*D*,*^, and we choose the sign by

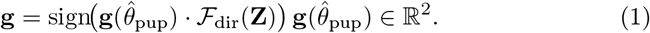

#### Refraction-aware Ellipse Correction

A refracted pupil ellipse *ℰ*_pup_ can be computed via a closed-form refraction operator ℛ given full 3D eyeball geometry, which is unavailable at inference [18,5] (Fig. 2I(d)). We therefore approximate the non-linear refraction mapping with a lightweight corrector *ℱ*_ref_, implemented as a 3-layer MLP (hidden dimension 128) operating on projection geometry *𝒫*^2*D*^ (Fig. 2I(c-i)):

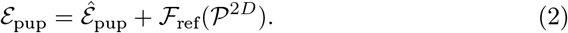

The corrector is pretrained on synthetic data and subsequently frozen during end-to-end training to preserve a stable refraction mapping (Fig. 2I(c-ii)).

#### Geometry-constrained Eye Reconstruction

Given the refracted ellipse *ℰ*_pup_ and projection geometry *𝒫*^2*D*^, eyeball parameters *𝒫* are recovered via ellipse-to-circle lifting. Let *Π*_*f,r*_(*ℰ*, **g**) → {**c, n**, *r*} denote the closed-form unprojection of an ellipse to a 3D circle under focal length *f*, assumed radius *r*, and prior direction **g** [15,17]. The pupil and iris ellipses are lifted as:

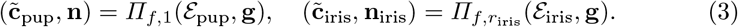

The recovered normal **n** defines the optic-axis-based gaze direction. Assuming concentric pupil and iris circles, the corrected pupil center **c**_pup_ is obtained by projecting 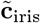 onto the line 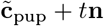. Given anatomical priors *𝒜*, the eyeball center **c**_eye_ and pupil radius *r*_pup_ are computed analytically (Fig. 2I(e)) [17,5].

Because reconstruction enforces geometric consistency, it compensates for inaccurate iris annotations via ellipse-to-circle alignment (Fig. 2II(a)).

#### Temporal Anatomical Stabilization

Frame-wise reconstruction is sensitive to segmentation noise and occlusion. We therefore impose sequence-level regularization by refining the eyeball center across a mini-batch. A lightweight temporal module ℱ_tem_ aggregates temporal tokens and geometric descriptors using a single-layer Transformer encoder (embedding dim. 128, 4 heads, feed-forward dim. 256), followed by a bounded MLP head predicting an anatomically constrained displacement:

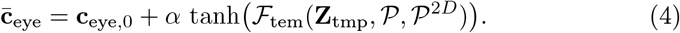

Here **c**_eye,0_ is a confidence-weighted prior. Assuming limited anatomical variation within a mini-batch, the refined center 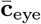 is shared across frames (Fig. 2I(f)). The optic axis and pupil radius are recomputed analytically, and gaze angles (*θ*_*H*_, *θ*_*V*_) are obtained by converting the optic axis to spherical coordinates.

### 2.2 Loss Function

The model is trained end-to-end using:

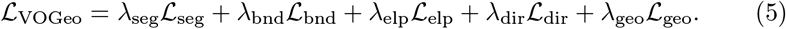

*ℒ*_seg_ combines region, boundary and smoothness constraints following [12]. *ℒ*_bnd_ enforces physiological constraints by penalizing pupil radius and gaze estimates outside predefined anatomical bounds. *ℒ*_elp_ is used for VOGeo-Gaze training and the refraction corrector *ℱ*_ref_ pretraining; it combines a parameter term (active only when training *ℱ*_ref_) and contour consistency minimizing algebraic distance. The direction loss minimizes angular discrepancies:

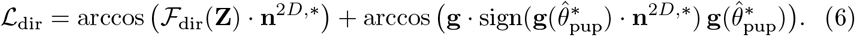

#### Geometry Loss

Let 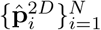 be uniformly sampled points on the predicted entrance pupil ellipse 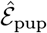 with 2D center 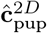. Each point defines a camera ray that intersects the cornea and refracts onto the 3D pupil circle. The refraction operator *ℛ*_*K,𝒜*_ computes refracted contour points 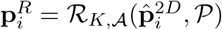 and a refracted pupil circle center 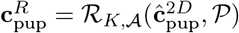 under intrinsics *K* and anatomical priors. We constrain refracted points to form a circle orthogonal to the optic axis (Fig. 2II(b)):

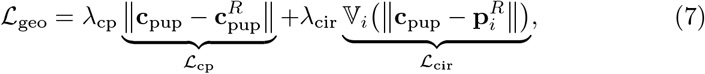

where the two terms correspond to center consistency and circularity.

## 3 Experiments and Results

### 3.1 Datasets and Experimental Setup

#### Training Data

VOGeo-Gaze was trained on the TEyeD dataset [7] using a subject-independent 8:1:1 train/validation/test split. Training samples were constructed using a sliding temporal window of *T* = 16 consecutive frames with stride 2, forming input tensors of size *B* × *T* × *C* × *H* × *W*. Frames were resized to 240 × 320 pixels and augmented with random zoom and horizontal/vertical flips. Sequences shorter than *T* were padded by repeating the last frame. Data were stored in LMDB format for efficient sequential access. The segmentation backbone was initialized from pretrained 3DeepVOG weights and fine-tuned jointly.

#### Synthetic Pretraining

The corneal refraction corrector *ℱ*_ref_ was pretrained on over 10 million synthetically generated eyeball configurations under fixed camera intrinsics *K* and anatomical priors *𝒜*. Eyeball parameters *𝒫*^*^ were randomly sampled within ranges (Table 1) yielding ground-truth refracted pupil ellipses 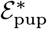, iris ellipses 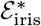, and prior gaze directions **g**\*. Entrance pupil ellipses 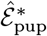 were generated using physics-based inverse ray tracing [3]. The network learned 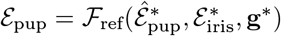 supervised by 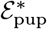 (Fig. 2I(c-ii)).

**Table 1.**
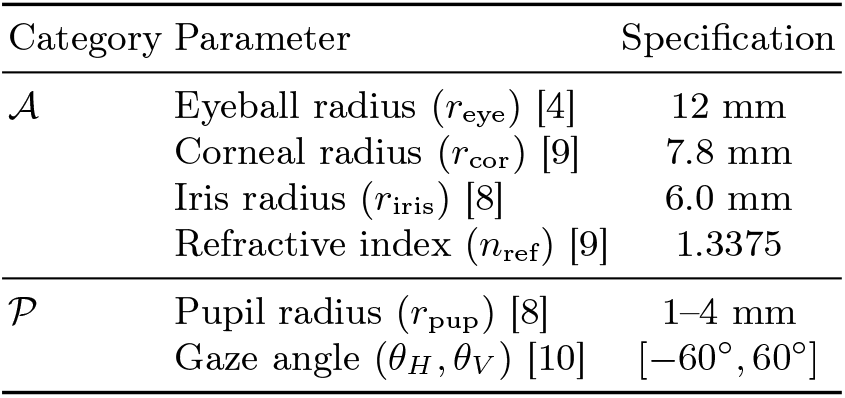
Anatomical priors 𝒜 and physiological parameter ranges 𝒫 used for geometric modeling and synthetic data generation.

#### Clinical Evaluation

Evaluation was conducted on 116 clinical VOG recordings from 17 patients and 19 healthy subjects acquired at LMU University Hospital Munich using the EyeSeeCam system. Protocols included caloric testing, video head impulse testing (vHIT), and oculomotor tasks (saccades, smooth pursuit, optokinetic nystagmus). All recordings were obtained with known camera intrinsic parameters and calibration sequences to create the necessary ground truth. The study was approved by the Ethics Committee of the Medical Faculty, Ludwig Maximilian University Munich, approval number 24-0108. Table 2 summarizes all datasets used in this study.

**Table 2.** Summary of datasets used in this study for model training, refraction-module pretraining, and clinical evaluation.

| Dataset | Size | Label | Usage |
| --- | --- | --- | --- |
| TEyeD [7] | 20M+ frames | Seg + gaze | Training |
| Synthetic Refraction | 10M+ samples | Refracted ellipses | $\mathcal{F}_{\text{ref}}$ pretrain |
| Clinical (EyeSeeCam) | 116 videos | Device gaze + ellipse | Evaluation |

#### Metrics and Implementation

Angular accuracy was measured using mean absolute error (MAE) after linear alignment [20]. Geometric consistency was quantified via the refraction-aware loss *ℒ* _geo_ (7), decomposed into *ℒ* _cp_ and *ℒ*_cir_. The model was implemented in PyTorch (Python 3.11) and trained on an NVIDIA RTX 4090 using AdamW (lr = 1 × 10^*−*5^, *β* = (0.9, 0.99), weight decay = 1 × 10^*−*4^) with cosine scheduling and 5-epoch linear warmup.

### 3.2 Comparison with Representative Paradigms

We compared VOGeo-Gaze against two paradigms: (A) Analytical geometry reconstruction (3DeepVOG [20], IrisFit [17,7]) and (B) label-driven learning (NVGaze [11], implemented via AcomoEye-NN [1], and CVL [14]). All Type B baselines were retrained on TEyeD with identical preprocessing. De^2^Gaze [19] was omitted as no public implementation is available, precluding fair reproduction.

#### Quantitative Results

As shown in Table 3, 3DeepVOG achieves the lowest MAE (0.29°/0.27°) with calibration and known intrinsics, while VOGeo-Gaze closely matches this performance (0.33°/0.35°) without either. VOGeo-Gaze is the only method besides 3DeepVOG exceeding 90% of recordings below 1° error in both components. Regression-based and non-refraction-aware methods show substantially higher error and dispersion, indicating the benefit of explicit refraction modeling. Moreover, label-driven approaches depend directly on dataset-provided gaze annotations, which may constrain achievable clinical accuracy. VOGeo-Gaze requires 9.2 GFLOPs, substantially lower than CVL (22.6 GFLOPs), enabling real-time processing (>300 FPS).

**Table 3.**
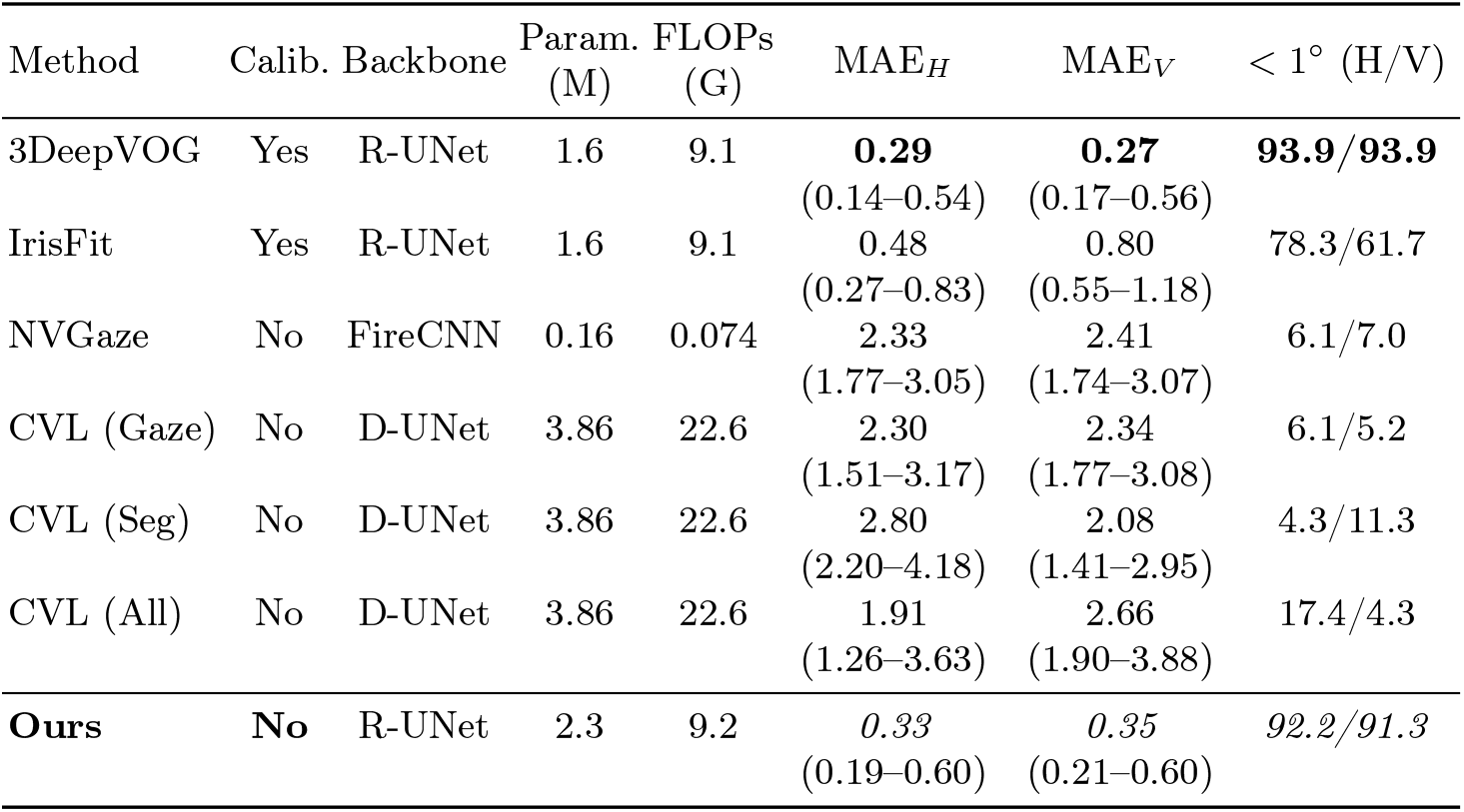
Benchmark on clinical VOG recordings (*N* = 116). MAE is reported as median (Q1–Q3) in degrees after alignment [20]. < 1° (H/V) indicates the percentage of recordings below 1° MAE (bold/italic font: best/second-best method).

| Method | Calib. | Backbone | Param.<br>(M) | FLOPs<br>(G) | MAE<sub>H</sub> | MAE<sub>V</sub> | $< 1^\circ$ (H/V) |
| --- | --- | --- | --- | --- | --- | --- | --- |
| 3DeepVOG | Yes | R-UNet | 1.6 | 9.1 | **0.29**<br>(0.14–0.54) | **0.27**<br>(0.17–0.56) | **93.9/93.9** |
| IrisFit | Yes | R-UNet | 1.6 | 9.1 | 0.48<br>(0.27–0.83) | 0.80<br>(0.55–1.18) | 78.3/61.7 |
| NVGaze | No | FireCNN | 0.16 | 0.074 | 2.33<br>(1.77–3.05) | 2.41<br>(1.74–3.07) | 6.1/7.0 |
| CVL (Gaze) | No | D-UNet | 3.86 | 22.6 | 2.30<br>(1.51–3.17) | 2.34<br>(1.77–3.08) | 6.1/5.2 |
| CVL (Seg) | No | D-UNet | 3.86 | 22.6 | 2.80<br>(2.20–4.18) | 2.08<br>(1.41–2.95) | 4.3/11.3 |
| CVL (All) | No | D-UNet | 3.86 | 22.6 | 1.91<br>(1.26–3.63) | 2.66<br>(1.90–3.88) | 17.4/4.3 |
| **Ours** | **No** | R-UNet | 2.3 | 9.2 | *0.33*<br>(0.19–0.60) | *0.35*<br>(0.21–0.60) | *92.2/91.3* |

#### Geometric Consistency and Qualitative Analysis

We used 3Deep-VOG as a calibrated analytical reference to evaluate geometric consistency via center consistency (*ℒ*_cp_) and circularity (*ℒ*_cir_). Paired Wilcoxon tests (*N* = 116) showed no significant difference for *ℒ*_cp_ (*p* = 0.17) or *ℒ*_cir_ (*p* = 0.73), indicating comparable geometric consistency.

Fig. 3 further illustrates failure cases of the calibration-based baseline under poor calibration or incorrect camera intrinsics, leading to erroneous gaze direction and inconsistent parameter estimation. In contrast, VOGeo-Gaze remains robust without relying on calibration and camera intrinsics.

**Fig. 3.**
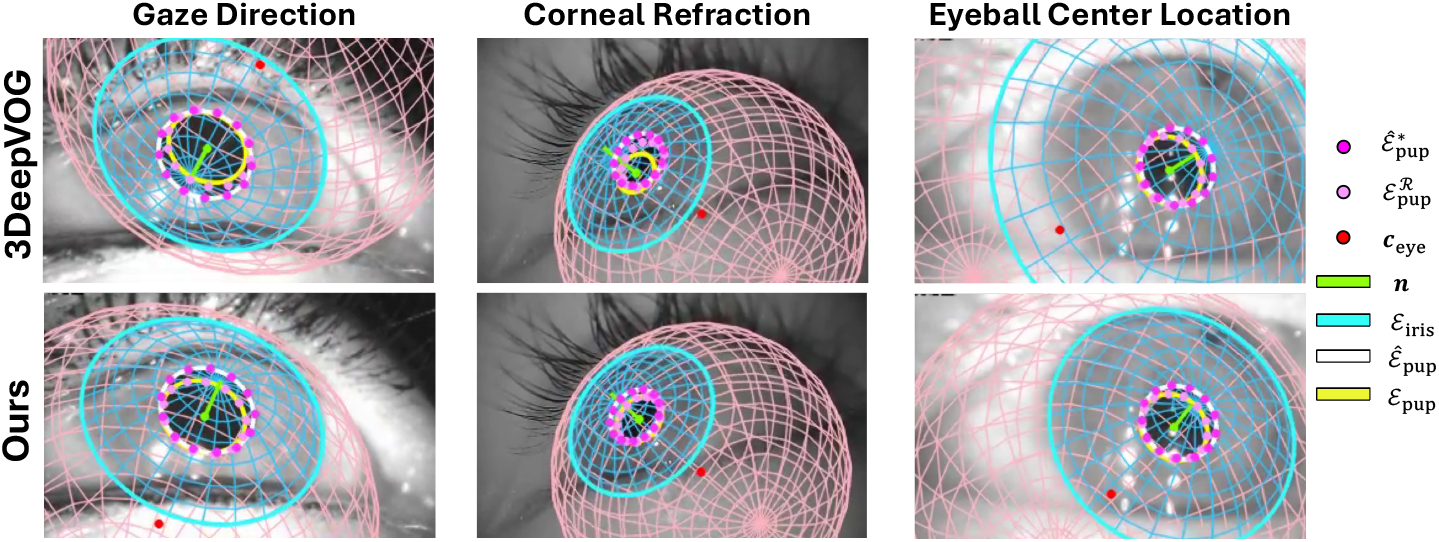
Qualitative comparison of analytical geometry reconstruction method (3Deep-VOG) and VOGeo-Gaze under poor calibration or incorrect camera intrinsics.

### 3.3 Ablation Study

Table 4 evaluates the impact of key components. Removing the corneal refraction corrector *ℱ*_ref_ leads to the largest performance drop, indicating that explicit refraction modeling is critical for sub-degree accuracy. Disabling the temporal refiner *ℱ*_tem_ moderately increases error, suggesting that temporal aggregation mainly improves stability. Training the full network from scratch (no pretrained segmentation and *ℱ*_ref_) degrades performance, highlighting the benefit of staged training. Finally, jointly fine-tuning *ℱ*_ref_ without freezing also reduces accuracy, indicating that maintaining a stable refraction mapping during VOGeo-Gaze training improves consistency.

**Table 4.** Ablation study on clinical VOG recordings (*N* = 116). Values are median (Q1–Q3) MAE in degrees after alignment.

| Variant | MAE<sub>H</sub> ↓ | MAE<sub>V</sub> ↓ | < 1° (H/V) ↑ |
| --- | --- | --- | --- |
| **Full (Ours)** | **0.33** (0.19–0.60) | **0.35** (0.21–0.60) | **92.2/91.3** |
| No $\mathcal{F}_{\text{ref}}$ | 1.62 (1.36–2.09) | 1.78 (1.45–2.22) | 61.5/56.8 |
| No $\mathcal{F}_{\text{ref}}$ freezing | 1.44 (0.85–1.82) | 1.50 (0.89–1.98) | 74.3/72.6 |
| No $\mathcal{F}_{\text{tem}}$ | 0.91 (0.44–1.77) | 0.98 (0.48–1.86) | 81.2/79.3 |
| No pretraining | 0.49 (0.29–0.92) | 0.55 (0.32–0.98) | 80.9/78.3 |

## 4 Conclusion

We presented VOGeo-Gaze, a calibration-free, refraction-aware framework for clinical gaze estimation that reconstructs anatomically interpretable eye parameters from segmentation-driven image cues. By explicitly modeling corneal refraction and enforcing geometric consistency, VOGeo-Gaze achieves sub-degree accuracy on 116 clinical VOG recordings without subject-specific calibration, camera intrinsics, or accurate gaze labels, closely matching 3DeepVOG. Future work will address adaptive estimation of anatomical priors, evaluate generalization to additional devices and public benchmarks, and extend the framework to broader calibration-free near-eye settings (e.g., VR/AR).

## Data Availability

Code and training data are available online

https://github.com/DSGZ-MotionLab/VOGeo-Gaze

https://es-cloud.cs.uni-tuebingen.de/d/8e2ab8c3fdd444e1a135/?p=%2FTEyeDS&mode=list

## Acknowledgments

This work was supported by the German Space Agency (DLR) on behalf of the Federal Ministry of Economics and Energy (Grant No. 50WB2236) and by the German Federal Ministry of Education and Research (Grant No. 13GW0490B).

## References

1. AcomoEye-NN: NVGaze gaze estimation with upgrades. https://github.com/czero69/acomoeye-NN, last accessed 2026/05/23

2. Agrawal, Y., Schubert, M.C., Migliaccio, A.A., Zee, D.S., Schneider, E., Lehnen, N., Carey, J.P.: Evaluation of quantitative head impulse testing using search coils versus video-oculography in older individuals. Otology & Neurotology 35(2), 283–288 (2014). 10.1097/MAO.0b013e3182995227

3. Aguirre, G.K.: A model of the entrance pupil of the human eye. Scientific Reports 9(1), 9360 (2019). 10.1038/s41598-019-45827-3

4. Bekerman, I., Gottlieb, P., Vaiman, M.: Variations in eyeball diameters of healthy adults. Journal of Ophthalmology 2014, Article ID 503645 (2014). 10.1155/2014/503645

5. Dierkes, K., Kassner, M., Bulling, A.: A novel approach to single camera, glint-free 3D eye model fitting including corneal refraction. In: Proceedings of the 2018 ACM Symposium on Eye Tracking Research and Applications (ETRA 2018), pp. 1–9. ACM, New York (2018). 10.1145/3204493.3204525

6. Dierkes, K., Kassner, M., Bulling, A.: A fast approach to refraction-aware eye-model fitting and gaze prediction. In: Proceedings of the 11th ACM Symposium on Eye Tracking Research and Applications (ETRA 2019), pp. 1–9. ACM, New York (2019). 10.1145/3314111.3319819

7. Fuhl, W., Kasneci, G., Kasneci, E.: TEyeD: Over 20 million real-world eye images with pupil, eyelid, and iris 2D and 3D segmentations, 2D and 3D landmarks, 3D eyeball, gaze vector, and eye movement types. In: 2021 IEEE International Symposium on Mixed and Augmented Reality (ISMAR), pp. 367–375. IEEE, Piscataway (2021). 10.1109/ISMAR52148.2021.00053

8. Gross, H. (ed.): Handbook of Optical Systems, Vol. 4: Survey of Optical Instruments. Wiley-VCH, Weinheim (2008). 10.1002/9783527623385

9. Guestrin, E.D., Eizenman, M.: General theory of remote gaze estimation using the pupil center and corneal reflections. IEEE Transactions on Biomedical Engineering 53(6), 1124–1133 (2006). 10.1109/TBME.2005.863952

10. Hansen, D.W., Ji, Q.: In the eye of the beholder: A survey of models for eyes and gaze. IEEE Transactions on Pattern Analysis and Machine Intelligence 32(3), 478–500 (2010). 10.1109/TPAMI.2009.30

11. Kim, J., Stengel, M., Majercik, A., De Mello, S., Dunn, D., Laine, S., McGuire, M., Luebke, D.: NVGaze: An anatomically-informed dataset for low-latency, near-eye gaze estimation. In: Proceedings of the 2019 CHI Conference on Human Factors in Computing Systems (CHI 2019), pp. 1–12. ACM, New York (2019). 10.1145/3290605.3300780

12. Kothari, R.S., Chaudhary, A.K., Bailey, R.J., Pelz, J.B., Diaz, G.J.: EllSeg: An ellipse segmentation framework for robust gaze tracking. IEEE Transactions on Visualization and Computer Graphics 27(5), 2757–2767 (2021). 10.1109/TVCG.2021.3067761

13. Myronenko, A.: 3D MRI brain tumor segmentation using autoencoder regularization. In: Crimi, A., Bakas, S. (eds.) Brainlesion: Glioma, Multiple Sclerosis, Stroke and Traumatic Brain Injuries. Lecture Notes in Computer Science, vol. 11384, pp. 311–320. Springer, Cham (2019). 10.1007/978-3-030-11726-9_28

14. Popovic, N., Christodoulou, D., Paudel, D.P., Wang, X., Van Gool, L.: Model-aware 3D eye gaze from weak and few-shot supervisions. In: 2023 IEEE International Symposium on Mixed and Augmented Reality Adjunct (ISMAR-Adjunct), pp. 746–751. IEEE, Piscataway (2023). 10.1109/ISMAR-Adjunct60411.2023.00161

15. Safaee-Rad, R., Tchoukanov, I., Smith, K.C., Benhabib, B.: Three-dimensional location estimation of circular features for machine vision. IEEE Transactions on Robotics and Automation 8(5), 624–640 (1992). 10.1109/70.163786

16. Schneider, E., Villgrattner, T., Vockeroth, J., Bartl, K., Kohlbecher, S., Bardins, S., Ulbrich, H., Brandt, T.: EyeSeeCam: An eye movement-driven head camera for the examination of natural visual exploration. Annals of the New York Academy of Sciences 1164, 461–467 (2009). 10.1111/j.1749-6632.2009.03858.x

17. Świrski, L., Dodgson, N.A.: A fully-automatic, temporal approach to single camera, glint-free 3D eye model fitting. In: Proceedings of the European Conference on Eye Movements (ECEM 2013), Lund, Sweden (2013).

18. Villanueva, A., Cerrolaza, J.J., Cabeza, R.: Geometry issues of gaze estimation. In: Advances in Human-Computer Interaction. InTech, Vienna (2008). 10.5772/5911

19. Xiao, Y., Bai, X., Chen, B., Su, H., He, H., Xie, L., Yin, E.: De2Gaze: Deformable and decoupled representation learning for 3D gaze estimation. In: Proceedings of the IEEE/CVF Conference on Computer Vision and Pattern Recognition (CVPR 2025), pp. 3091–3100. IEEE, Piscataway (2025). 10.1109/CVPR52734.2025.00294

20. Zhao, J., Ahmadi, S.-A., Decker, J., Möhwald, K., zu Eulenburg, P., Zwergal, A., Flanagin, V.L., Wuehr, M.: 3DeepVOG: An open-source framework for real-time, accurate 3D gaze tracking with deep learning. Digital Biomarkers 10(1), 21–31 (2026). 10.1159/000549948

